# Perceptions of partial gland ablation for prostate cancer among men on active surveillance: A qualitative study

**DOI:** 10.1101/2020.09.15.20195388

**Authors:** Sonia S. Hur, Michael Tzeng, Eliza Cricco-Lizza, Miko Yu, Jessica Ancker, Erika Abramson, Christopher Saigal, Ashley Ross, Jim C. Hu

## Abstract

**Objectives:** Partial gland ablation (PGA) therapy is an emerging treatment modality that targets specific areas of biopsy proven prostate cancer (PCa) to minimize treatment-related morbidity by sparing benign prostate. This qualitative study aims to explore and characterize perceptions and attitudes toward PGA in men with very-low-risk, low-risk, and favorable intermediate-risk PCa on active surveillance (AS).

**Design:** 92 men diagnosed with very-low-risk, low-risk, and favorable intermediate-risk PCa on AS were invited to participate in semi-structured telephone interviews on PGA.

**Setting:** Single tertiary care center located in New York City.

**Participants:** 20 men with very-low-risk, low-risk, and favorable intermediate-risk PCa on AS participated in the interviews.

**Main outcome measures:** Emerging themes on perceptions and attitudes toward PGA were developed from transcripts inductively coded and analyzed under standardized methodology.

**Results:** Four themes were derived from twenty interviews that represent the primary considerations in treatment decision-making: (1) the feeling of psychological safety associated with low risk disease; (2) preference for minimally invasive treatments; (3) the central role of the physician; (4) and the pursuit of treatment options that align with disease severity. Eleven men (55%) expressed interest in pursuing PGA only if their cancer were to progress, while 9 men (45%) expressed interest at the current moment.

**Conclusions:** Though an emerging treatment modality, patients were broadly accepting of PGA for PCa with men primarily debating the risks versus benefits of proactively treating low-risk disease. Additional research on men’s preferences and attitudes toward PGA will further guide counseling and shared decision-making for PGA.

**Key Messages:** *What is already known about this subject?:* Partial gland ablation (PGA) is an emerging treatment option for prostate cancer (PCa) that allows for the targeting of areas of biopsy proven PCa with the goal of minimizing treatment-related morbidity by avoiding treatment of non-cancerous areas of the prostate. We sought to explore patients’ beliefs and attitudes toward partial gland ablation.

*What are the new findings?:* We define treatment attributes that are significant to men with localized PCa, and one important theme is of the treatment intensity matching the severity of the disease. PGA appeals to men as middle ground that encompasses the duality of curative treatment and preservation of quality of life.

*How might these results affect future research or surgical practice?:* The exploratory themes need further exploration and may be incorporated into shared decision-making discussions in men with low-risk disease, as PGA emerges as a treatment option.

## Introduction

Partial gland ablation (PGA) is an emerging modality in the treatment of prostate cancer (PCa) that allows for the targeting of areas of biopsy proven PCa with the goal of minimizing treatment-related morbidity. The European Association of Urology recognizes PGA as an investigational modality, and most published studies focus on its role in men with very-low-risk to intermediate-risk PCa. [1,2] Despite its potential, consensus on patient selection for PGA is lacking, as is intermediate to long-term outcomes on cancer control. [2–4]

Studies of PGA for prostate cancer have been shown to have very low rates of complications that require medical intervention, incontinence, and erectile dysfunction, respectively. It has potential as the least minimally invasive alternative treatment to traditional whole gland treatments such as surgery or radiation therapy. [2–4] However, post-treatment prostate specific antigen (PSA) surveillance is unreliable with PGA due to background PSA levels produced by the untreated prostate. As such, monitoring with prostate biopsy is the most definitive surveillance approach for PGA; [5] however, prostate biopsy has risks of pain, bleeding and infection. Moreover, high-level evidence for PGA is lacking, and men face uncertainty over cancer progression when pursuing non-definitive treatment. [6] A recent multi-specialty consensus conference spearheaded by the U.S. Food and Drug Administration (FDA) outlined a paradigm to evaluate PGA through a prospective randomized trial with men on active surveillance (AS) as a comparator. [5] While the majority of urologist believe that PGA will become a standard option, [7] patient attitudes and perceptions toward PGA have yet to be defined.

Currently, PGA is most commonly offered to men with low-risk to intermediate-risk PCa. [8] Men with low-risk disease pursue AS due to evidence that Grade Group 1 cancers are often indolent in nature. [6] Men with favorable intermediate-risk prostate cancer are higher risk for progression on surveillance but may also consider this management approach. [9] Active surveillance, i.e. monitoring with curative intent, avoids the risk of side effects such as urinary incontinence and erectile dysfunction associated with whole gland treatments, such as radical prostatectomy and radiation therapy. [10] Surveillance can however take a psychological toll and men on AS have reported the need to develop coping mechanisms related to anxiety over the long-standing chronicity of untreated disease. [11] These accounts of patient experience on AS suggest there may be a role for PGA for some of these men.

In the shared decision-making process for PCa management, appropriate choice of treatment modality requires a comprehensive understanding of patient values and beliefs. To our knowledge, there is currently little evidence regarding men’s attitudes and perspectives toward PGA. Therefore the goal of our study was to conduct interviews with men on AS to elucidate their consideration of treatment options and describe their opinions of PGA.

## Subjects and Methods

### Approach

The constant comparative method as described by Glaser and Strauss was used for thematic analysis. [12] In this approach, data is first inductively coded and analyzed to identify concepts. As concepts evolve, data is revisited and undergoes focused coding in an iterative process to develop themes. Interviews were conducted in a semi-structured fashion to allow for the emergence of new patterns in data. Thematic saturation was initially observed after 15 interviews, when no new themes were identified. Interviews were conducted past the threshold of thematic saturation and concluded at 20 patients.

### Participants

We identified English-speaking men on active surveillance at Weill Cornell Medicine with very-low-risk, low-risk, and favorable intermediate-risk PCa who had been seen in clinic at least once during the past 2 years. All participants are currently on AS for very-low-risk, low-risk or favorable intermediate-risk PCa consistent with National Comprehensive Cancer Network guidelines. [13]

### Recruitment and data collection

Ethical approval was obtained from the Weill Cornell Institutional Review Board and all participants provided written informed consent. Eligible patients were mailed letters of invitation to participate in the study and offered to be interviewed by mail or phone. Patients who did not respond were contacted by phone also, when possible. Two research assistants uninvolved with medical care, conducted all interviews over the phone at the convenience of the participants.

Prior to the interview segment on PGA, participants were provided a short summary of recent evidence-based findings as described by the European Association of Urology in 2018. [2] As most patients had never heard of PGA before, this provided necessary context for discussion. Information was presented as objectively as possible. The mean duration of interviews was 32.1 (standard deviation 8.0) minutes. Relevant medical information, including age, years since diagnosis, serum prostate specific antigen, and Gleason score, was extracted from patient medical records after interviews.

The first 10 interviews were conducted with one researcher leading the interview and both researchers present at all times. These interviews served as a training period to promote standardization of process. The final 10 interviews were then conducted separately. All interviews were digitally recorded. The final iteration of the interview template is shown in **Figure 1**.

**Figure 1.**
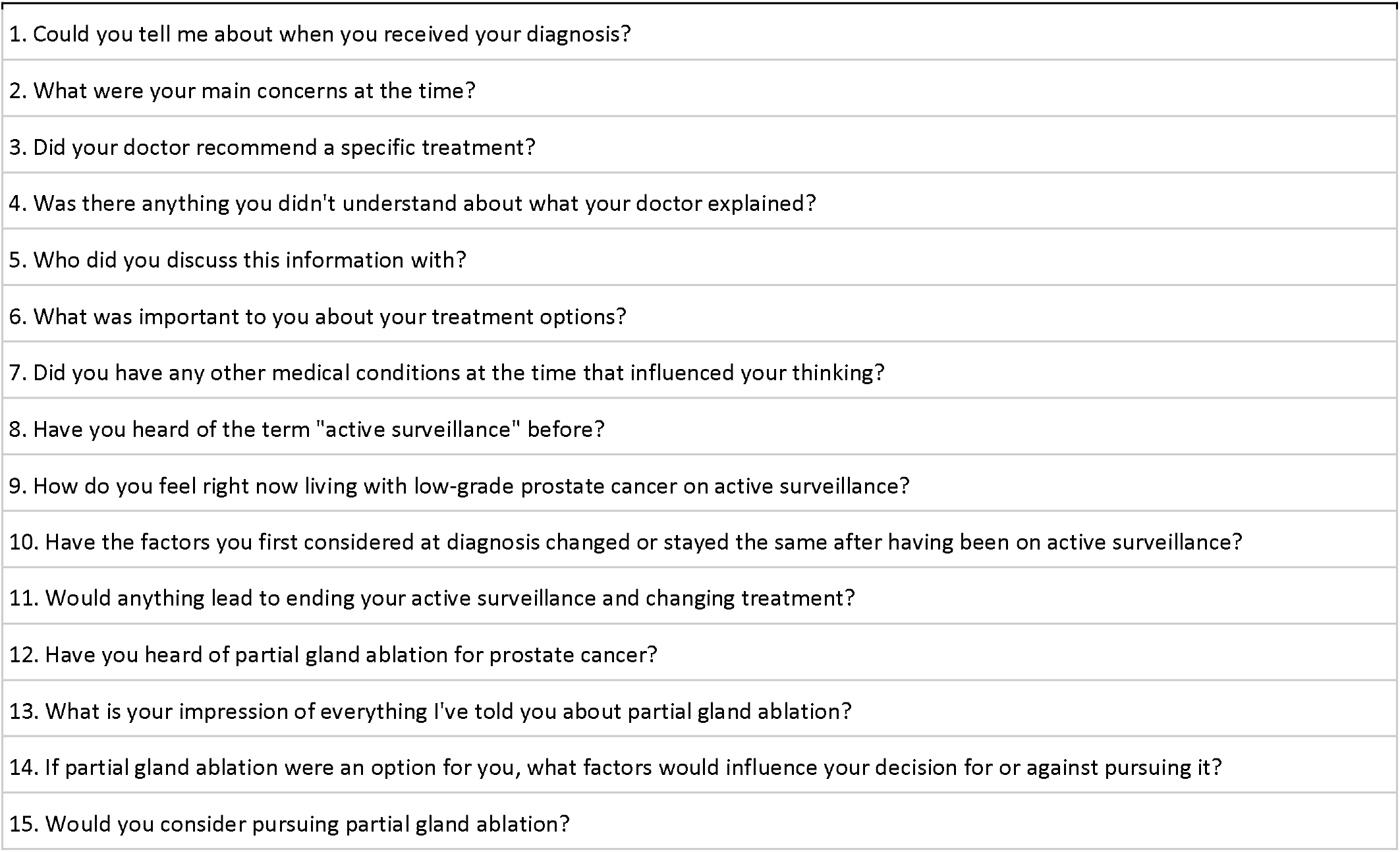
Interview template.

### Analysis

Interviews were transcribed, inductively coded and analyzed. Every five interviews, transcripts were reviewed with senior researchers to discuss concepts, perform focused coding, and develop themes. The interview template was then edited to reflect these emerging themes. Research assistants inductively coded and analyzed the first 10 transcripts together to standardize methodology and negotiate emerging themes. The final 10 transcripts were inductively coded and analyzed individually. Microsoft Excel was used for data management.

## Results

Out of 92 total men contacted, 20 consented for participation and were interviewed. Patient demographics and clinical characteristics are shown in **Table 1**. Eighty-five percent of subjects were white. Ninety-five percent of patients completed college or attained graduate degrees, and 75% had annual household incomes exceeding $110,000. 6 (30%) participants were diagnosed with very low-risk, 12 (60%) with low-risk PCa, and 2 (10%) with favorable intermediate-risk PCa. Four themes pertaining to men’s pursuit of treatment options were identified: (1) the perception of psychological safety in the diagnosis of low-risk prostate cancer;(2) the prioritization of minimally invasive options that have fewer side effects; (3) the dependence on the provider in the decision-making process; (4) and matching the aggressiveness of treatment with the degree of disease severity (**Figure 2**). Of the 20 men interviewed, 11 (55%) men expressed interest in pursuing PGA only if their cancer were to progress, while 9 (45%) men expressed interest at the current moment.

**Table 1.** Participant demographics and clinical characteristics.

| Table 1. Participant demographics and clinical characteristics |  |
| --- | --- |
| Median (IQR) age, years | 66 (62.3-71.5) |
| Median (IQR) years since diagnosis | 1.9 (0.9-3.7) |
| Median (IQR) PSA, ng/mL | 5.9 (4.2-7.5) |
| Gleason score, n (%) |  |
| 3+3 | 18 (90) |
| 3+4 | 2 (10) |
| Race/ethnicity, n (%) |  |
| White | 17 (85) |
| Black | 2 (10) |
| Asian | 1 (5) |
| Educational attainment, n (%) |  |
| High school degree | 1 (5) |
| Bachelor’s degree | 10 (50) |
| Master’s degree | 2 (10) |
| Doctoral degree | 7 (35) |
| Household income, n (%) |  |
| 60k-110k | 5 (25) |
| >110k | 15 (70) |
| N/A | 1 (5) |
| Employment, n (%) |  |
| Full-time | 15 (75) |
| Part-time | 1 (5) |
| Retired | 3 (15) |
| Unemployed | 1 (5) |
| Marital status, n (%) |  |
| Single | 1 (5) |
| Married | 18 (90) |
| Divorced | 1 (5) |

**Figure 2.**
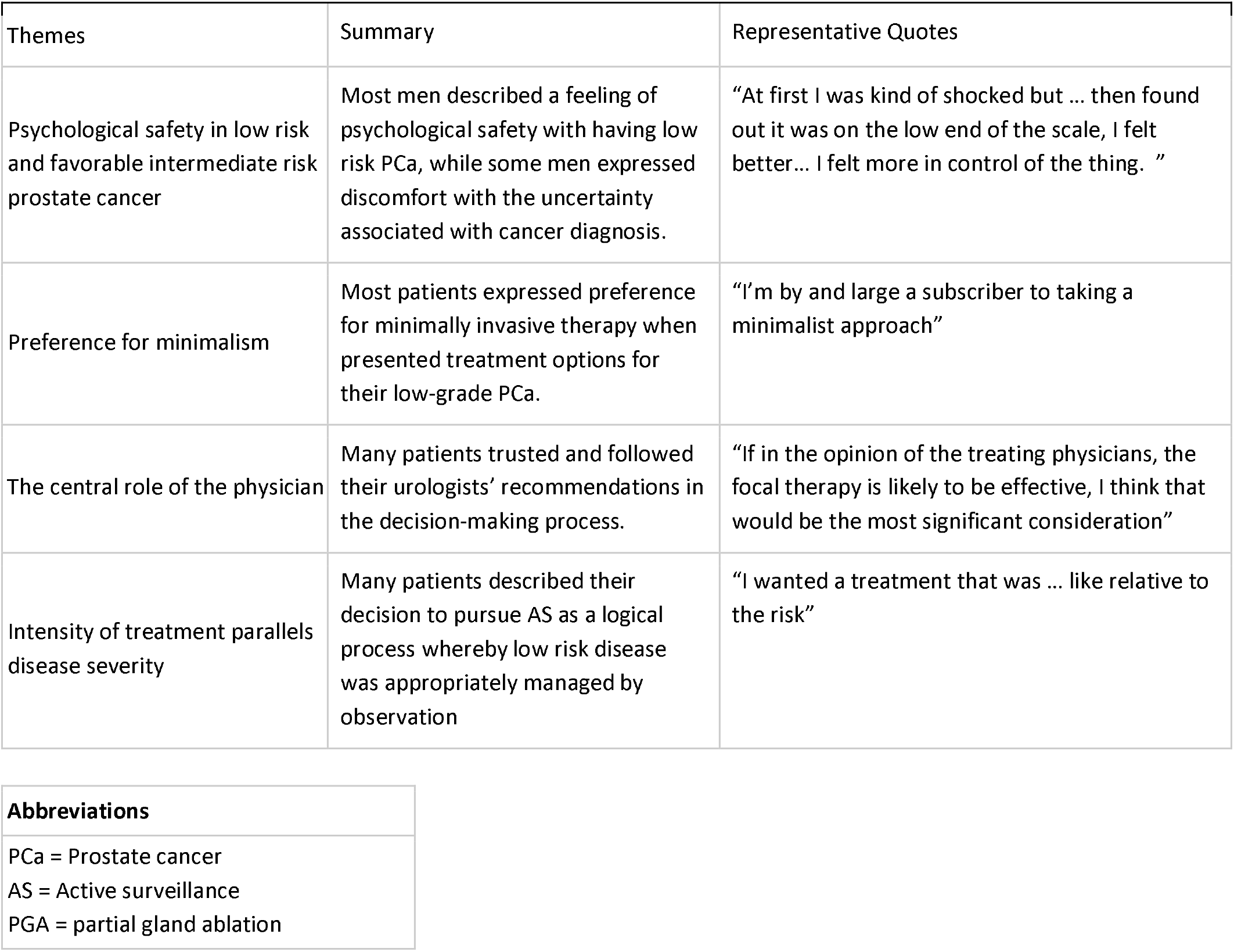
Themes related to men’s perceptions of partial gland ablation.

### Psychological safety in low-risk and favorable intermediate-risk prostate cancer

Most men perceived their cancer as low risk, allowing them to find comfort in pursuing AS over definitive treatment. Although some men were first concerned at diagnosis, most came to understand their cancer as a chronic condition that they “needed to follow-up on,” but not immediately treat because there wasn’t a “huge chance of it spreading.” One man acknowledged an “element of uncertainty” being on AS but didn’t feel the need to “dwell on” it. Moreover, one man claimed that his PCa “was the easiest thing I’ve had to deal with” among all his other medical conditions. Many men also perceived their cancer as non-fatal, commonly quoting that they were “more likely to die with it than of it.” The ubiquity of cancer was often identified by multiple patients, with one man citing that he heard “if you’re 70 you have a 70% chance of having PCa. Eighty is 80%.” One quote from a 60-year-old man exemplifies the lack of urgency most patients felt about their diagnosis:

> I really don’t think about it very much. To me, it’s sort of on par with my hypertension… Hypertension, pre-diabetes, the prostate--and to me they’re on a similar level… I think of them all on kind of the same… low level of health concern that I need to be careful about and watch and do the best I can.

Discussions regarding PGA for patients accepting of AS yielded mixed results. While some embodied the idea of “just the fact that it exists does not make me want to go and get it,” others expressed interest in further discussions at their next appointment.

Although most patients were satisfied with their current management, two particular men demonstrated a large psychological toll associated with their cancer. One 59-year-old patient whose parent died from cancer described PCa as “this thing hanging over your head” and likened AS to “check[ing] in with the parole officer.” Another 67-year-old man described AS as “playing with fire” and “cutting corners.” His thought process below illustrates his feeling of brokenness associated with having PCa:

> I really don’t care how painful the procedure is. If they want to take a piece out, then take the piece out. If they blast me with radiation, that’s what they gotta do… I don’t feel like dying from cancer. That’s been my only concern… I want it fixed… I don’t want me walking around with cancer between my legs.

In these patients who were considering discontinuation of AS, PGA seemed “interesting” and something they wanted to discuss more with their physician.

### Preference for minimalism

Most patients expressed preference for minimally invasive therapy when presented treatment options for their low grade PCa. Some men had an innate inclination towards a less invasive procedure without any definite reason. One 56-year-old patient explained, “there is no scientific explanation… or no deep explanation” for wanting “to save a part of my body that can continue its’ function in my body.”

For most men, the inclination towards the less invasive treatment reflected their desire to avoid side effects associated with surgical removal of the prostate. Most patients understood radical prostatectomy to be a “potentially life-changing procedure”, naming loss of sexual and urinary functions as main reasons. Two of the youngest patients, in their mid 50s, detailed desire to avoid “impotence” or “wearing a diaper.” For one 70-year-old man, the potential side effects were more worrisome than the uncertainty of living with cancer.

> I was more worried about the consequences of surgery—the possible consequences-than… the actual diagnosis of cancer… the possibility of incontinence or… reduced or lower sexual activity.

Another patient echoed this sentiment towards PGA, stating “I would be willing to live with that uncertainty in order to avoid the side effects and invasiveness of the radical surgery.” Many patients equated minimally invasive treatment with less side-effects. For example, one man stated if treatment, including PGA, “is less invasive, it wouldn’t have many side effects” and “will involve less risk for complication.” Most men thought PGA was a safer alternative to radical prostatectomy, and voiced plans to discuss with their urologist.

However, not all men found the minimally invasive option of PGA as intuitive. These men prioritized the treatment of cancer more important than preservation of their urinary and sexual function. One 57-year-old patient reflected on his initial confusion before choosing AS as his cancer management: “the only thing that can happen is it can get worse, why don’t we just do the procedure and get rid of it?” One patient, 68-year-old, compared choosing between curative definitive treatment and possible urinary incontinence as having to pick his “poison”, but found that “it’s still better than ending up dead.” Three men perceived PGA as a “band-aid” or a “temporary fix” as opposed to radical prostatectomy because of the remaining uncertainty after focal treatment.

Additionally, approximately one fourth of men interviewed touched on the invasiveness and frequency of periodic prostate biopsies while on AS. A 71-year-old man worried that prostate biopsies came with their own risk of sepsis and possible antibiotic resistance. Another 73-year-old patient reported opting out of annual prostate biopsies in favor of non-invasive interventions with a naturopath. However, most men did not recognize prostate biopsies as a new disadvantage when transitioning to PGA from AS. One 57-year-old man explained:

> The negative of it is something that I’ll have to do with active surveillance anyway. I still have to be monitored, I still have to do my biopsy, now I still have to do all of that. So… the negative part of it hasn’t changed what I’m current doing.

While one patient expressed continued concern over prostate biopsies in regard to PGA, most men expressed sustained interest in learning more about PGA.

### The central role of the physician

Many patients trusted and followed their urologists’ recommendations in the decision-making process. Most men found confidence in physicians who were “up-to-date” and “major”, or renowned, in their specialty. Some men placed a high value in their physician’s proficiency in the procedures they perform, stating “I have looked for the best surgeon to have the best outcome… regardless of the method or procedure I choose.” Another patient highlighted his physician-patient relationship as reason for his trust, “I feel very comfortable with him and as long as I’m following the procedures that he told me to, I’m fine.” Moreover, three patients completely absolved themselves of personal research to rely on their physician’s recommendations:

> If my doctor says that’s the best option, then I’ll follow my doctor… If the doctor says yes, I’m not going to question him.

Most patients actively pursued information beyond the urology visit, usually online. Some men recounted anecdotes from peers with history of PCa, while a few turned to scientific studies. Many patients consulted with their friends and family, though most said this had no actual influence on their decision. Some patients referred to their spouses as a “sounding board, someone [they] can bounce ideas off of” or “more of a supporting role [to] calm [them] down.”

Most men asserted complete understanding of their disease and treatment options, as elucidated by their urologist. Patients believed their physicians were acting in their best interest, especially with the recommendation of a “non-invasive” and “less aggressive” approach. In addition, men found their urologist’s recommendation of AS reinforced their appreciation of the low risk nature of their condition and lessened the psychological distress associated with the cancer diagnosis. Furthermore, some men were encouraged by their primary urologists to pursue second opinions at a “teaching hospital and seek out a younger doctor.” Few patients reported their optimistic genomic biopsy results reassuring them into AS. Men described a collaborative relationship with their urologists but felt that they were ultimately in control of the decision:

> No, my doctor and I made the decision. My family played, you know, a secondary. I make the decisions about my life; they don’t. If the doctor says it is time for us to… need a decision, at that time, I will decide what needs to be done.

In considering PGA, men were even more inclined to depend on their urologists’ recommendations and expertise, given the “experimental” nature of PGA.

> My surgeon’s confidence in it and his history of performing each of these procedures. I would only feel confident if he has a long history and has done it 800 times or so.

However, not all patients found their specialists to be candid in their treatment approach, stating “nobody really recommended any other treatment options other than the ones that they liked… or they performed.” These patients felt the need to “inform myself of the different therapies that were open to me.”

### Intensity of treatment parallels disease severity

Many patients described their decision to pursue AS as a logical process whereby low-risk disease was appropriately managed by observation. AS was described as “an apt measure based on the diagnosis” compared to the “over-reactive invasive” option of radical prostatectomy. As one patient explained, “I just felt like it was the right thing to do. It made sense to me. It was logical… as long as there’s… regular and consistent monitoring… you know, it seemed that’s what I should do.” The lack of symptoms of low-risk disease also played a role in deciding against definitive treatment in a patient whose follow-up biopsy was negative: “If my prostate is working… If I’m not having any of the symptoms or I’m not having any discomfort… Why would I go and do anything radical?” One patient who also saw a naturopath for his PCa even felt that the management process of AS itself was excessive:

> It’s our culture that’s leading us to—let’s do this procedure let’s do this test and that test and more… what is the bottom-line improvement in one’s life when we’re, you know, talking about something that’s not that aggressive.

As a result, many patients expressed that they would proceed to surgery or radiation only if it were “necessary” or if the cancer were “worsening,” which they felt were determined by the combination of test results and provider recommendation.

This step-wise approach was expressed similarly in discussions regarding PGA, although with varied outcomes. Eleven men (55%) expressed interest in pursuing PGA only if their cancer were to progress, while 9 men (45%) expressed interest at the current moment. In considering further discussion on PGA with their provider, some expressed interest only if future biopsies showed worse outcomes, since “the treatment would be aggressive to match the more aggressive diagnosis.” A few examples are illustrated:

> I’m actually happy with the active surveillance. Someone would have to say that ‘your risk has increased’ and if there were something that said ‘your risk has increase’ then I would prefer to have something like focal therapy or focal surgery than a full excision of the prostate. I think that I would need a really bad Gleason score to opt in for some kind of treatment. It’s gotta be a real black and white issue.

Yet, others felt that the relatively conservative and localized approach of PGA deserved consideration and at least a discussion with their provider, even at their current risk category.

> If he thinks he can get rid of what little I have by doing this treatment, I definitely have an interest in it.
>
> The finding was in a very small area. And that the Gleason score was not highly aggressive. So therefore… it would make sense to me to do the focal therapy as opposed to doing something more radical.
>
> I would think that given the ability to focus on the areas of concern, just those parts of the prostate that… might suggest cancer could be targeted fairly well. You know, I’d be game for something like that.

## Discussion

Our study identifies four key thought processes in considering PGA among men with very-low-risk, low-risk, and favorable intermediate-risk PCa on active surveillance. Namely, these include the perception of psychological safety in a lower risk diagnosis, the prioritization of minimally invasive options with fewer side effects, the dependence on the provider in the decision-making process, and the consideration of treatment aggressiveness with respect to disease severity. These concepts were further explored in the context of considering PGA as a treatment option, for incorporation into shared decision-making discussions. Our cohort reflects recent treatment expansion of AS to include men with favorable intermediate-risk PCa on AS along with men with very-low-risk and low-risk PCa on AS. [5] Overall, 9 patients expressed interest in speaking with their provider to discuss PGA at their current risk category and 11 expressed interest in the event their cancer progresses.

Most men described a feeling of psychological safety with having low-risk disease on AS. This finding parallels of other studies that identified men’s acute perception of the low-risk category and non-immediate threat to life. [10,14] However, some men in our study expressed a significant discomfort with the uncertainty of their diagnosis and were considering other treatment options. This uncertainty has been observed by others and shown to be associated with lower quality of life. [15] Men expressing uncertainty about AS were more receptive to PGA, and many patients comfortable on AS were also interested in further discussion at their current risk level.

Earlier studies of AS assert that patients with low-risk PCa valued prolonged survival over preservation of urinary and sexual functions, [16,17] thus are likely to pursue definitive treatment over AS. [18,19] However, our findings concur with more recent findings that indicate a cultivating preference for minimally invasive treatments by men with a refined understanding of their low-risk condition. [10,20] Currently it is widely known that men with low-risk PCa managed on AS have higher quality of life, [21] explaining men’s motivation to trade off low uncertainty over survival for preservation of urinary function. [20] PGA was appealing as a curative alternative to AS and a less invasive option to radical prostatectomy for men in this study. However, fifteen percent of men perceived PGA as a temporary cure when compared to whole gland treatments as the uncertainty of recurrence remains.

Trust in the physicians and their recommendations has been shown to play a key role in pursuing any treatment strategy, [6,22,23] although some men also value playing an active role [24] and taking ownership over their choices. [10,23,25] Regardless, greater patient knowledge is also associated with greater decision-making difficulty, [26] which suggests an important role in shared decision-making with providers. Our study is consistent with prior findings that patients who established a collaborative relationship with their physicians [27] are more likely to seek their physician’s expertise and ‘decisional support’ [25] when surveying new treatment options, including PGA. However, few patients did not find the recommendations for focal treatments to be candid, as they found specialists to be predisposed to recommending procedures they mainly perform.

Many men identified a logical process in their decision-making in which their low-risk disease was appropriately managed by the observational nature of AS. Previous studies have identified men’s understanding of the rationale behind AS as a contributing factor towards staying on AS. [14] Our findings contribute that men on AS respect the escalation of treatment invasiveness that parallels disease progression. This novel concept of treatment intensity matching disease can be explored in shared decision-making discussions with the introduction of PGA as an intermediate option on the prostate cancer treatment spectrum. Interestingly, these men differed in their opinions on where PGA stands in this hierarchy, with some finding it an appropriate option for low-risk disease and others believing it only necessary for higher risk disease.

Earlier studies found men with low-risk PCa having higher overall satisfaction with care with definitive treatments such as radical prostatectomy or radiotherapy than with AS. [27] However, definitive treatments for patients with localized PCa is currently considered overtreatment, [18] introducing unnecessary side effects. Nevertheless, many men elect to undergo definitive therapy to address the uncertainty of cancer. [16] Although short-term oncological outcomes vary and may depend on the specific modality used, [2,3,28] PGA emerges as middle ground that encompasses the satisfaction of curative treatment and preservation of quality of life. [18]

There are multiple limitations to our study. First, this was a qualitative study of a small sample of 20 men on AS from a single tertiary care center located in New York City. Compared to the general population of prostate cancer patients, patients in this study were mostly white men who had attained Baccalaureate degrees or higher, residing in areas of higher income. Our findings may not be generalizable to men from different backgrounds. Sampling bias must be considered given that our cohort includes only men who volunteered to be interviewed. Second, the time since diagnosis averaged 2.6 years in our study and may have contribute to recall bias, loss of details about the initial consultation, and establishment of comfort regarding their diagnosis and treatment options. Additionally, semi-structured interviews required probing patients with unprompted questions, adding to both interviewer and response bias. Lastly, phone interviews do not provide visual cues and may contribute to the loss of nonverbal data and contextual information and misinterpretation of responses. [29] However, phone interviews offer facial anonymity, which may empower patients to disclose sensitive information more readily. [29]

In conclusion, as PGA develops there has been an emerging consensus that men with tumor characteristics eligible for AS may the best candidates for PGA; however, there is little knowledge of men’s attitudes and perspectives on PGA. Herein we identify four themes and their relation to men’s considerations of PGA: the feeling of psychological safety associated with low-risk disease, a preference for minimally invasive treatment, the central role of the physician, and the pursuit of treatment option intensity that parallels disease severity. In a highly educated sample that is experienced with AS with low-risk PCa, we demonstrate that almost half of men have potential interest in PGA, despite low grade evidence concerning intermediate and long-term outcomes.

## Data Availability

All data relevant to the study are included in the article.

## Acknowledgement

Jim Hu is supported by The Frederick J. and Theresa Dow Wallace Fund of the New York Community Trust.

